# Attitudes of People Living with Dementia and their Carers towards the use of Generative Artificial Intelligence to inform Structured Medication Reviews

**DOI:** 10.64898/2026.06.24.26356103

**Authors:** Bethany Linder, Jiamin Du, Nuno Tavares, Tingting Zhu, Prayag Tiwari, Sundus Jawad, Anna Elizabeth Seeley, Subhashisa Swain, Jaheeda Gangannagaripalli

## Abstract

**Background:** Polypharmacy is common in people living with dementia (PLwD) and associated with adverse outcomes. Although Structured Medication Reviews (SMRs) are recommended to optimise medication regimens, their delivery is often constrained by limited healthcare resources. Artificial intelligence (AI) may support SMRs, yet little is known about how it is perceived by PLwD and their carers. This study explored their experiences of polypharmacy, views on SMRs, and attitudes towards use of AI tools in SMRs.

**Methods:** Semi-structured interviews with 12 PLwD experiencing polypharmacy and two focus groups with 14 carers were conducted via Microsoft Teams or telephone and analysed using Reflexive Thematic Analysis.

**Results:** Two themes were constructed: experiences of SMRs, and attitudes towards AI in SMRs. Participants described challenges in managing polypharmacy, with carers often playing a central role in supporting adherence and monitoring side effects. Experiences of SMRs varied widely. SMRs were most valued when clinicians were empathetic and able to offer personalised guidance. Participants viewed AI use in SMRs positively, provided that such tools were well validated and used to assist rather than replace healthcare professionals. AI was viewed as having the potential to reduce administrative burden and support more person-centred care. However, some had concerns regarding patient safety and data security, highlighting the need for appropriate regulation and human oversight.

**Conclusion:** Participants were supportive of AI use in SMRs, despite concerns about safety, data security and disclosure of AI use, and emphasised the importance of patient–clinician interactions and lived experience involvement in AI tool development.

## Introduction

Polypharmacy is defined as the use of five or more medications[1], which can lead to adverse outcomes, including adverse drug events, falls, hospitalisations, and mortality[2,3]. People living with dementia (PLwD) are more likely to be affected by co-morbidities and polypharmacy compared to the general population[4,5]. Though polypharmacy is necessary for some patients, finding strategies to prevent inappropriate polypharmacy (such as use of multiple medications that are no longer clinically appropriate and may cause harm) and overprescribing in this population is crucial, as this could improve patient safety and quality of life of PLwD[6]. PLwD and their carers experience disproportionately high and complex medication burden, requiring more coordinated, person-centred medication management support[7]. Structured medication reviews provide important opportunities to optimise medication use and reduce the overall burden associated with medicines.

Structured medication reviews (SMRs) were introduced in the UK in 2020 to address inappropriate polypharmacy in the National Health Service (NHS)[8]. SMRs involve a healthcare professional reviewing a patient’s prescriptions and medical records to optimise the regimen to ensure it is safe and meets the patient’s needs[8]. A secondary goal is to deprescribe where possible to minimise potential drug interactions and side effects, and prevent waste through eliminating unnecessary medications[8]. National Institute for Health and Care Excellence (NICE) guidelines emphasise the importance of SMRs being grounded in shared decision making, with healthcare professionals taking the best available evidence and the patient’s preferences into account when optimising a patient’s medication[9]. Though estimates vary, research suggests that only up to half of eligible patients in the UK receive a yearly SMR[10,11], potentially due to time constraints and lack of resources in primary care[12,13]. Constraints can be exacerbated in the context of PLwD, because dementia is a complex condition and is associated with co-morbidities that require more time to explore[14,15]. Furthermore, communication difficulties may make gathering patient history and preferences particularly challenging[16].

Artificial intelligence (AI) technology could offer an important strategy to aid clinicians in conducting SMRs and ultimately enable a greater number of patients to receive SMRs[17]. The data processing capabilities of generative AI models could reduce the time clinicians spend sieving through numerous healthcare records, letters and test results[18]. In addition, these tools may be used to identify individuals at risk of polypharmacy, highlight futile treatment, and improve medicine management[19,20]. This information can directly inform clinicians’ decisions regarding medication optimisation in individuals living with polypharmacy, especially people living with polypharmacy and dementia. However, attitudes towards using AI technology in healthcare and the needs of people affected by dementia concerning SMRs are not well understood. This study aims to explore the challenges of managing medications for PLwD, the concerns and priorities regarding SMRs, and the perspectives of PLwD and carers around using AI to support conducting SMRs.

## Methods

### Study Design

This study adopted a qualitative design underpinned by an interpretive theoretical position[21]. Semi-structured interviews and focus groups were used as these methods allowed for flexibility in discussions.

### Participants and Recruitment

Participants comprised PLwD and informal carers of PLwD residing in the United Kingdom. Inclusion criteria for PLwD included people aged over 18 years and living with a diagnosis of early-stage dementia and polypharmacy (≥5 medications). Meanwhile, carers were recruited if they provided unpaid care to someone with a diagnosis of dementia and polypharmacy.

Carers were recruited through the National Institute for Health and Care Research (NIHR)’s *People in Research* service, and PLwD were recruited through advertisements in the newsletters of four support services for PLwD (two national, one based in Northwest England, and one based in Southeast England). Eligible individuals who responded to the advertisements were recruited using a convenience sampling approach.

### Study Materials and Data Collection

Carers took part in two focus groups (with seven participants in each group) lasting approximately 1.5 hours and PLwD participated in individual interviews lasting up to 60 minutes. Although face-to-face options were offered, all participants elected to take part remotely through Microsoft Teams or telephone calls. Both focus groups and interviews followed the same semi-structured interview guide (Supplementary File 1). Participants were asked about their experiences with managing medication and SMRs, and thoughts about using AI in SMRs. To support participants’ understanding, a brief description of an AI-based medication review tool was provided to all participants, which included intended purpose, types of data it uses, and how it supports clinical decision-making. The interview guide was informed by two workshops on the use of AI during SMRs for patients with dementia conducted with ten healthcare professionals of various healthcare backgrounds and Patient and Public Involvement and Engagement (PPIE) representatives.

The focus groups were facilitated by BL as lead interviewer, supported by JD or JG as co-facilitators. Individual interviews were conducted by BL. All interviews and focus groups were carried out between September and November 2025. Interviews and focus groups were transcribed verbatim by the interviewer, with any identifying information removed during transcription. No participants had met the interviewers prior to the study, and no follow-up interviews took place.

### Ethics

The study received a favourable ethical opinion from the University of Oxford Medical Sciences Interdivisional Research Ethics Committee (MS IDREC) (ref: 788094). Participants provided either written or verbal informed consent prior to participation. Optional pre-interview calls were offered to familiarise participants with the process, discuss any accessibility needs and identify possible adjustments (e.g., having a shorter interview).

Participants were assigned participant numbers and identifiable information was removed from quotes to maintain confidentiality. All sessions were recorded and stored securely on a password-protected university server at the University of Oxford.

### Data Analysis

Data were analysed using reflexive thematic analysis[22]. An inductive approach was used, allowing themes to be constructed from the data rather than from pre-existing frameworks. Focus groups and interviews transcripts were analysed together to identify shared themes. To ensure rigour, BL, JD and JG independently analysed three transcripts and kept reflexive diaries, then met to compare their initial interpretations. BL coded the remaining transcripts and constructed initial themes. The three authors then met repeatedly to develop, refine and verify themes, ensuring each was supported by sufficient data before finalisation.

## Results

### Participant Characteristics

Twenty-six participants (12 PLwD and 14 carers) participated in the study. Participant characteristics are presented in Table 1.

**Table 1.** Characteristics of study participants.

| Characteristic | PLwD (*n* = 12) | Carers (*n* = 14) |
| --- | --- | --- |
| **Age group, n (%)** |  |  |
| 18–40 | 0 (0.0) | 7 (50.0) |
| 41–50 | 0 (0.0) | 3 (21.4) |
| 51–60 | 1 (8.3) | 3 (21.4) |
| 61–70 | 4 (33.3) | 0 (0.0) |
| 71–80 | 6 (50.0) | 1 (7.1) |
| Prefer not to say | 1 (8.3) | 0 (0.0) |
| **Gender, n (%)** |  |  |
| Man | 8 (66.7) | 11 (78.6) |
| Woman | 3 (25.0) | 3 (21.4) |
| Prefer not to say | 1 (8.3) | 0 (0.0) |
| **Ethnicity, n (%)** |  |  |
| Black | 0 (0.0) | 8 (57.1) |
| Asian | 0 (0.0) | 3 (21.4) |
| Mixed | 0 (0.0) | 2 (14.3) |
| White | 11 (91.7) | 1 (7.1) |
| Prefer not to say | 1 (8.3) | 0 (0.0) |
| **Employment status, n (%)** |  |  |
| Full time | 0 (0.0) | 1 (7.1) |
| Part time | 0 (0.0) | 11 (78.6) |
| Unemployed | 0 (0.0) | 1 (7.1) |
| Retired | 11 (91.7) | 1 (7.1) |
| Prefer not to say | 1 (8.3) | 0 (0.0) |
| **Geographical area, n (%)** |  |  |
| East Anglia | 0 (0.0) | 1 (7.1) |
| East Midlands | 1 (8.3) | 0 (0.0) |
| London | 0 (0.0) | 1 (7.1) |
| North-east England | 0 (0.0) | 1 (7.1) |
| North-west England | 4 (33.3) | 5 (35.7) |
| Northern Ireland | 1 (8.3) | 1 (7.1) |
| Scotland | 1 (8.3) | 0 (0.0) |
| South-east England | 2 (16.7) | 3 (21.4) |
| West Midlands | 1 (8.3) | 2 (14.3) |
| Yorkshire and the Humber | 1 (8.3) | 0 (0.0) |
| Prefer not to say | 1 (8.3) | 0 (0.0) |
| **Type of dementia, n (%)** |  |  |
| Alzheimer's disease | 1 (8.3) | 4 (28.6) |
| Dementia with Lewy Bodies | 2 (16.7) | 0 (0.0) |
| Frontotemporal dementia | 3 (25.0) | 1 (7.1) |
| Mixed | 1 (8.3) | 2 (14.3) |
| Vascular dementia | 3 (25.0) | 5 (35.7) |
| Unknown | 1 (8.3) | 2 (14.3) |
| Prefer not to say | 1 (8.3) | 0 (0.0) |

### Qualitative findings

Two themes were constructed; (1) Perceived pitfalls of SMRs, and (2) Attitudes Towards AI in SMRs. Each theme emerged from the combination of several subthemes (See Table 2 for a summary of these themes and subthemes). Participant quotations are labelled according to participant number and group, with participants P1–P14 representing carers and participants P15–P26 representing PLwD.

**Table 2.**
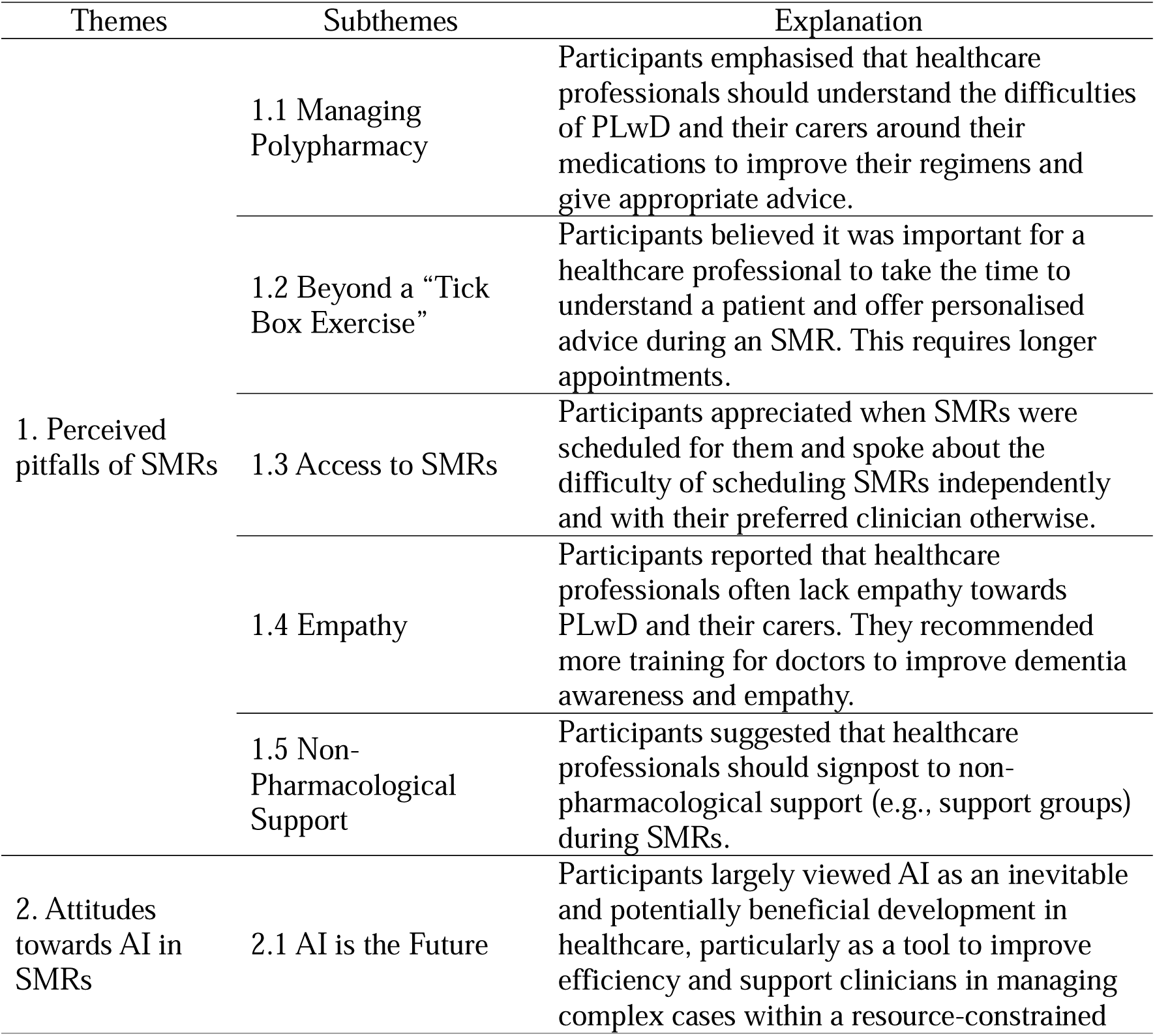
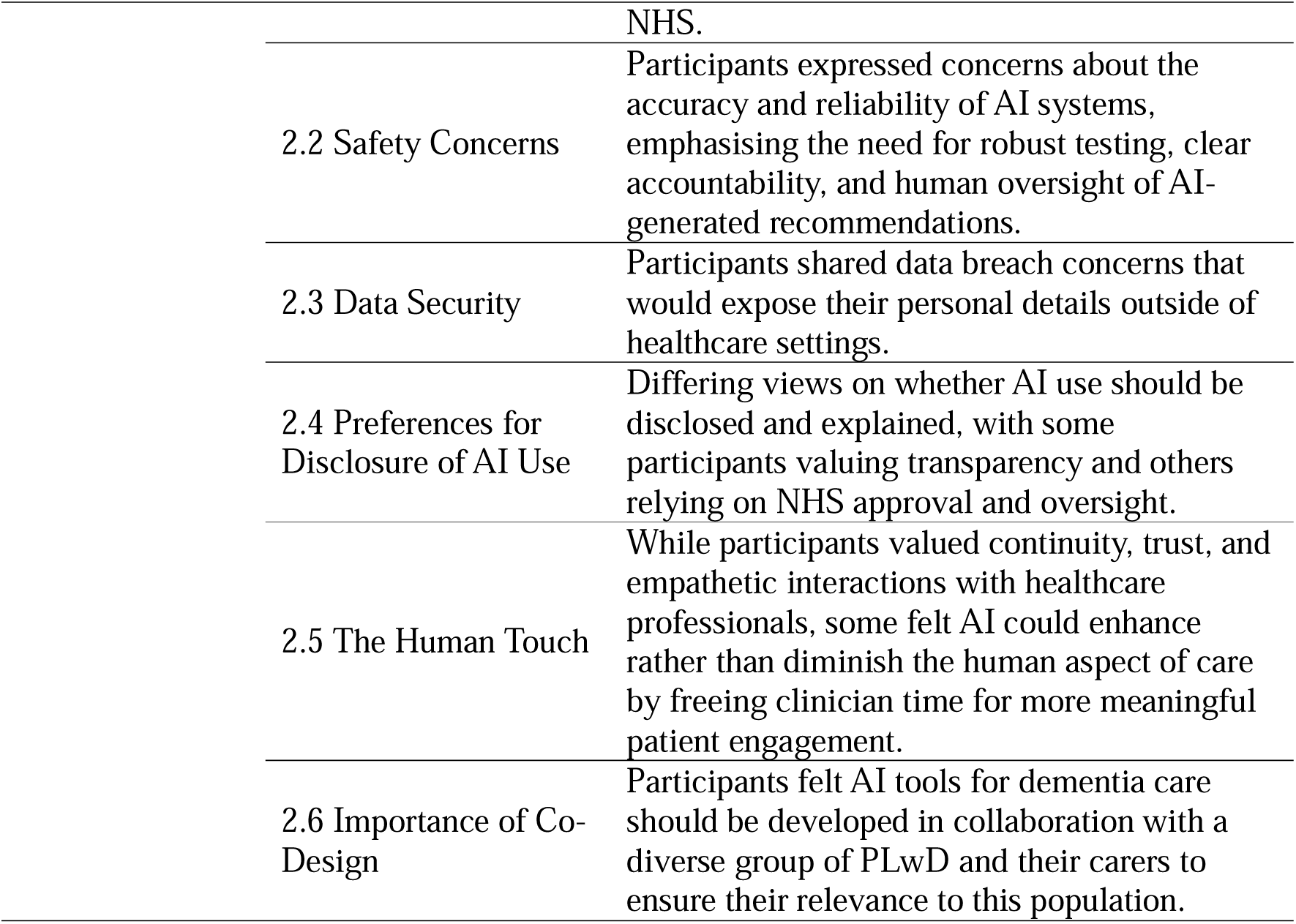
Overview of themes and subthemes derived from qualitative data.

| Themes | Subthemes | Explanation |
| --- | --- | --- |
| 1. Perceived pitfalls of SMRs | 1.1 Managing Polypharmacy | Participants emphasised that healthcare professionals should understand the difficulties of PLwD and their carers around their medications to improve their regimens and give appropriate advice. |
|  | 1.2 Beyond a “Tick Box Exercise” | Participants believed it was important for a healthcare professional to take the time to understand a patient and offer personalised advice during an SMR. This requires longer appointments. |
|  | 1.3 Access to SMRs | Participants appreciated when SMRs were scheduled for them and spoke about the difficulty of scheduling SMRs independently and with their preferred clinician otherwise. |
|  | 1.4 Empathy | Participants reported that healthcare professionals often lack empathy towards PLwD and their carers. They recommended more training for doctors to improve dementia awareness and empathy. |
|  | 1.5 Non-Pharmacological Support | Participants suggested that healthcare professionals should signpost to non-pharmacological support (e.g., support groups) during SMRs. |
| 2. Attitudes towards AI in SMRs | 2.1 AI is the Future | Participants largely viewed AI as an inevitable and potentially beneficial development in healthcare, particularly as a tool to improve efficiency and support clinicians in managing complex cases within a resource-constrained |

|  |  |
| --- | --- |
|  | NHS. |
| 2.2 Safety Concerns | Participants expressed concerns about the accuracy and reliability of AI systems, emphasising the need for robust testing, clear accountability, and human oversight of AI-generated recommendations. |
| 2.3 Data Security | Participants shared data breach concerns that would expose their personal details outside of healthcare settings. |
| 2.4 Preferences for Disclosure of AI Use | Differing views on whether AI use should be disclosed and explained, with some participants valuing transparency and others relying on NHS approval and oversight. |
| 2.5 The Human Touch | While participants valued continuity, trust, and empathetic interactions with healthcare professionals, some felt AI could enhance rather than diminish the human aspect of care by freeing clinician time for more meaningful patient engagement. |
| 2.6 Importance of Co-Design | Participants felt AI tools for dementia care should be developed in collaboration with a diverse group of PLwD and their carers to ensure their relevance to this population. |

### Theme One: Perceived pitfalls of SMRs

Participants reported varied experiences with SMRs. Some participants had regular SMRs which they found very helpful, and some considered SMRs *“a waste of time”* (P15, PLwD), and some had never been offered an SMR at all. This theme will report on the valued components of an SMR and factors that reduced their usefulness to people affected by dementia.

### Managing Polypharmacy

*“I can’t take them myself anymore because I either drop them, or I forget.” (P22, PLwD)*

Adhering to medication regimens was described as becoming increasingly difficult as dementia progressed. PLwD required more support to manage their medication over time, through setting reminders on apps, keeping medication diaries, or help from carers. Some described how it was becoming more difficult to understand whether unexplained symptoms were side effects of new medication or a part of dementia progression, because some PLwD can lack insight into aspects of their health.

> *“I don’t think I’ve had any side effects off any [medication]…but then, if I did, I would probably think maybe it’s something else. Because with my dementia…like if I was in pain, I wouldn’t recognise necessarily where that pain was coming from.”* (P22, PLwD)

Participants felt it was important that healthcare professionals were mindful of these challenges with medication, and valued advice about medication management and side effects during SMRs. Simplifying regimens during SMRs was also valued. The staff organising dosette boxes and ensuring that medication appearances (e.g. colour, shape, packaging) remain consistent can help PLwD to remember which medications they have taken and improve compliance.

> *“For me, challenges have been around when the packages are changed, colours of tablets have been changed, size has been changed…then my relative always is suspicious and she says to me, ‘but I don’t usually take this one…I used to take the pink one.’”* (P13, carer)

### Beyond a Tick Box Exercise

*“In a 5-minute consultation, they didn’t have the time to actually investigate properly and do their job properly.” (P15, PLwD)*

Some participants believed that SMRs would not be helpful if conducted as “*tick box exercises”*. Instead, individuals suggested that healthcare professionals should follow a holistic approach and have more allocated time for SMRs. They felt that some healthcare professionals did not engage with them in a meaningful way and just read a list of questions about medications aloud. To some, this left the impression that the healthcare professionals were not interested in helping them and were just fulfilling a requirement.

> *“It’s another tick box exercise. They’ve got a job to do. They’ve got 3 minutes or whatever they allocate to this…I just don’t think that they take it seriously.”* (P17, PLwD)

Short and time-pressured appointments were perceived as a barrier to meaningful engagement, reducing opportunities for healthcare professionals to understand individual circumstances and deliver personalised advice. Participants emphasised the importance of having longer appointments to conduct SMRs meaningfully. This was particularly important for those who had communication difficulties and needed additional time to express themselves.

> *“I have the language variant of Frontotemporal [dementia]. So, I can get a bit blank when it comes to [finding the right words]…so, the appointments can be quite stressful.” (P26, PLwD)*

### Access to SMRs

*“It’s impossible to see a GP nowadays anyway. You can never get an appointment.” (P20, PLwD)*

Accessing appointments with healthcare professionals was described as challenging, and participants indicated that the process of scheduling an SMR should be simplified. Those who had SMRs scheduled for them automatically at regular intervals (e.g. every 6 months or year) valued the convenience of this.

> *“I know when [my SMR] is coming up…and I don’t have to make an appointment with anybody.”* (P17, PLwD)

It may be beneficial for healthcare professionals to do more to make patients aware of SMRs and how to access a review, as not all participants were made aware of SMRs when they were first diagnosed with dementia or first took on caring responsibilities.

> *“I think there should be some structure or at least information for patients and carers. To say if we haven’t called you, you can call us and book a medications review. If I hadn’t gone to this meeting, I don’t think I would have known [that SMRs exist].”* (P13, carer)

### Empathy

*“I don’t think [doctors] realise what it’s like to live with dementia” (P16, PLwD)*

A lack of empathy was present in some clinical encounters, with some healthcare professionals failing to adapt their communication style to meet the needs of PLwD (e.g., not speaking slowly or repeating information). Those who spoke with empathetic healthcare professionals valued the opportunity, as this helped make their SMRs more reassuring and useful. For example, some participants spoke about the importance of feeling *“listened to”* (P3, carer) during appointments and felt more confident that they were not taking any medication that could be harmful if the healthcare professional was more attentive during their SMR. Seeing the same healthcare professional and building up rapport over time was also seen as helpful in this regard.

> *“There’s none of that continuity, I don’t even know who my GP is anymore. So, it’s difficult to have that conversation with [someone you don’t know], and I found it a lot easier when it was somebody that knew me.”* (P26, PLwD)

The inclusion of carers in discussions was highlighted as important by both PLwD and carers, as their perspectives helped prevent important information from being overlooked. Yet some carers had been excluded and/or felt their input was not valued by healthcare professionals.

> *“I often feel like I’m treated as an extra rather than someone who’s living with this reality every day, and I feel like the review should recognise the carer’s role…and actually take our input into account when these decisions are being made.”* (P3, carer)

### Non-Pharmacological Support

*“There’s more to dementia and treating it than giving out drugs.” (P17, PLwD)*

Participants recommended that healthcare professionals should consider a patient’s situation *“holistically”* (P26, PLwD) and consider giving advice that can improve their health and wellbeing beyond the use of medication. Although non-pharmacological support does not form part of an SMR, participants felt it was important to be offered support beyond pharmacological strategies. This would include signposting to dementia organisations, referrals to mental health support, advice about coping strategies, and advice about maintaining a healthy lifestyle. Some felt like the importance of this type of advice was overlooked by healthcare professionals.

> *“[I want to be asked] ‘how are you coping?’ - it’s not so much just the medication, but it’s the coping mechanisms that you’re using…social prescribing these days is another thing that GPs do, isn’t it? And weight loss and…I think those sorts of things would be helpful [to discuss in an SMR].”* (P17, PLwD)

### Theme Two: Attitudes Towards AI in SMRs

When discussing the use of AI in SMRs, participants suggested that AI could potentially improve their experiences with SMRs. However, they also expressed some worries about the safety of using AI in healthcare and were concerned about the accuracy and safety of potential tools. Participants emphasised the importance of including people with lived experience of dementia in the development of AI tools for dementia care, as this can help to ensure the tool is person-centred.

### AI is the Future

*“I think AI is the future and we’re going to have to get used to it” (P21, PLwD)*

AI was frequently framed as *“the future”* (P21, PLwD) or simply as *“just progress”* (P23, PLwD), with its use in healthcare viewed as inevitable. Participants who used generative AI in their personal life felt particularly positive about the use of AI in healthcare, as they saw it as a tool that could help healthcare professionals provide more efficient and effective care.

> *“I’m all for it. It’s a shortcut. I mean, I don’t say that my doctor is not allowed to use a pocket calculator, do I?…Because a calculator is just a tool, and all AI is, is a tool, that’s all. To help you to produce a good result quicker.”* (P15, PLwD)

Some viewed the move towards AI as a necessary step to help the NHS save resources. For those who found it difficult to make an appointment with a healthcare professional, they would value an AI tool that could monitor patients and flag those in need of medical care.

> *“Money is getting tighter. People are just trying to do a lot more for a lot less money, where AI comes in, that can help in numerous ways.”* (P20, PLwD)

Others expressed concern about healthcare professionals becoming overly reliant on AI tools, as this could cause problems if the system was not working correctly. They emphasised that care should be taken before AI is implemented in healthcare.

> *“What happens if we put all of our eggs in that one basket and things don’t work? Can we go back? […] My worry is that, will those clinicians lose the ability to be able to carry out these types of assessments?”* (P19, PLwD)

### Safety Concerns

*“I’m concerned about using [AI] for something as serious as a medication review” (P2, carer)*

Concerns were raised about potential risks of AI for patient safety, by those who were aware that generative AI models may produce *“hallucinations”* (inaccurate information) and can be susceptible to biases. Participants were concerned about who would be liable if a patient was harmed due to recommendations that came from AI.

> *“It’s still quite early to, in my opinion, to use AI to make this kind of medical analysis, since there’s a tendency for AI models to hallucinate, most of the time…how do you ensure that the model doesn’t mix up patient’s data, you know?”* (P2, carer)

Therefore, participants expressed that requiring a healthcare professional to review and approve an AI tool’s recommendation would be reassuring.

> *“Probably my biggest concern is that the final say should never be left with the AI. It must always be superseded by a qualified person to make the final decisions.”* (P24, PLwD)

AI was framed as having the potential to enhance patient safety, particularly through its capacity to identify problems that may be overlooked in clinical review. This was particularly relevant for participants with multiple long-term conditions as they spoke about how healthcare professionals can find it difficult to take their complex needs into account.

> *“I’ve got what they call complex needs because I’ve got so many different conditions, and for a doctor to remember all that and be aware of all that [is difficult], so often they’re [only able discuss one condition at a time]. An AI, when that’s doing an assessment, it’s going to be taking all your conditions into consideration and all your medications, so, its potential is absolutely phenomenal because it could do a better job than the doctor.”* (P24, PLwD)

### Data Security

*“The only worry is if the NHS gets hacked, my details could go well, wherever! That’s a worry…I don’t want my details going around the world.” (P25, PLwD)*

The prospect of AI accessing medical records was attached to fears of heightened vulnerability to breaches or unauthorised access. Additionally, missing data in medical records were another concern, particularly if they have moved regions or records have not been added by secondary care teams. If medical records are incomplete or contain inaccurate information, this could impact the quality of an AI tool’s output.

> *“People’s records are not always correctly written up…it could well be that [AI’s] decisions have been made based on incorrect information…data gets lost, medical records get lost.”* (P17, PLwD)

Other participants were comfortable with their medical records being accessed if using an AI tool had the potential to improve their healthcare, particularly if they had confidence that the NHS could adequately protect their data.

> *“I guess in a strange kind of way, technology has a lot of information about us without us realising, even now. So, if the safety measures are put into place so that wrong people don’t get hold of that kind of information…if you’re going to gain more from it and the risks are limited, then it’s sort of got to be a good thing.”* (P26, PLwD)

### Preferences for Disclosure of AI Use

Participants had varying views about whether they would want their healthcare professional to inform them that they were using an AI tool and to explain how the tool worked. Some would feel reassured by understanding how and why the tool was used.

> *“I’d also be reassured if the GP takes that time to explain what the computers are showing them so that I can understand how it’s helping with the decision…that would really will win my trust.”* (P3, carer)

Others discussed how they do not have to expertise around AI or technology in healthcare and so would not understand how an AI programme worked. They did not feel it was necessary to be told about the use AI tools in healthcare because they had confidence that any tool approved for use in NHS would have been properly tested and safe to use.

> *“A system as big as the NHS would need regular checks. I know they have their own IT departments that do this, so I’d be more than happy for the GP to work off an NHS system.”* (P18, PLwD)

### The Human Touch

*“AI can do so much, but you still need the human touch at the end of it” (P18, PLwD)*

Some participants were concerned that the development of AI tools could lead to having less contact with healthcare professionals, or that AI could replace their doctor. They emphasised the benefits of having a good relationship with their doctor, as they have developed a strong understanding of their preferences and needs that an AI tool wouldn’t know about.

> *“I’ve been going [to my GP] now for 40 odd years, so I think he’s got used to me by now. He knows what I can take on, and what I can’t.”* (P18, PLwD)

Others felt their interactions with doctors had been rushed and they hadn’t been able to provide the *“human touch”* (P20, PLwD) they want. Some commented on how AI could facilitate person-centred care by creating summaries that help healthcare professionals gain an understanding of a patient and avoid some of the disruptions caused by a lack of continuity of care that were discussed in the previous theme. Furthermore, by AI taking on some of their tasks, this would free up the healthcare professional to interact more with a patient during an appointment

> *“Seeing the GP use the computer as a tool while also talking to us and listening to our input from treating us like people, like real people. I think that will also make us feel satisfied because it will just show that the technology is there to support the better care of it all, rather than just replacing the human connection, because we still have the human connection from the GP.”* (P3, carer)

Participants believed that effective use of AI in medication reviews required sensitivity to personal preferences, overall wellbeing, and lifestyle context in order to generate genuinely personalised guidance. As discussed previously in the ‘Non-Pharmacological Support’ sub-theme, participants reported that healthcare professionals did not always personalise their guidance during SMRs, and so AI could make a meaningful contribution in this way.

> *“AI with medication should be looking at the person’s physical and mental health, overall, general health and well-being…and make suggestions for healthier options and highlighting things you need to be aware of…that makes it more person-centred.”* (P26, PLwD)

### Importance of Co-Design

*“It’s about us and it should be for us.” (P6, carer)*

Participants emphasised that when developing technology aimed at improving the care of PLwD, it is important for researchers or developers to involve a diverse group of PLwD and carers in the design to ensure it is relevant, acceptable, and responsive to the needs of dementia.

> *“Have developers made it…dementia friendly? Have they taken into account the multifaceted needs of people and not just had a conversation with four people with Alzheimer’s? […] they really need to spend a lot of time with a wide range of people with dementia, to get a good working model.”* (P19, PLwD)

## Discussion

This study explored the experiences of PLwD and their carers of SMRs in the context of managing polypharmacy, and their attitudes around using AI to support SMRs. Overall, participants described facing significant challenges around medication management, variable experiences with SMRs, but shared positive attitudes towards the use of AI tools in SMRs, provided the tools could be proven safe, reliable, and effective.

Consistent with existing literature, participants reported substantial difficulties with managing complex medication regimens. For example, participants living with dementia had difficulties remembering to take their medication and were becoming more reliant on reminders from their carers over time [23,24] and had difficulty with understanding whether symptoms were attributable to medication side effects or dementia progression. Participants also highlighted challenges arising from changes in medication appearance, including alterations in packaging, tablet colour, and tablet size, consistent with findings from previous studies examining pill appearance changes[25,26]. These findings highlight the importance of healthcare professionals being mindful of these medication challenges when conducting SMRs for PLwD. Simplifying medication regimens, providing medication management aids such as dosette boxes, and minimising unnecessary changes in medication appearance may help support medication management in this population[25–28].

Varied experiences of SMRs mirror findings of previous studies which found the quality and perceived usefulness of SMRs vary based on how they were delivered[17]. SMRs that were perceived as useful involved an empathetic clinician who had the time to understand the participant’s situation and offer personalised guidance. Participants particularly valued continuity of care and building trust and rapport with a clinician over time[29,30].

Conversely, SMRs that were perceived as unhelpful were short, rushed, and often conducted by clinicians who lacked understanding or empathy around dementia. These findings align with previous literature around how time constraints were a significant barrier to offering high-quality SMRs [29,31] and how care can be improved by dementia awareness and empathy training for clinicians[32].

Participants reported inconsistent access, format, and time allocations for SMRs, with some never having had one. This also suggests that individuals with dementia and carers do not receive adequate or accessible information about the purpose of appointments with healthcare professionals[33]. Participants preferred when SMRs were scheduled for patients at a predictable interval (e.g., annually). This improves accessibility to primary care and timely diagnosis of health or medication issues for these individuals [34]. Finding strategies for improving the consistency and accessibility of SMRs is an important target for healthcare services and future research. Similar to previous research, participants in our study suggested that AI could be used to monitor patient records and identify and contact patients who are in need of an SMR and prompt them to make an appointment[35].

Overall, participants were receptive to the concept of using AI tools to support SMRs, particularly when the AI tool could be used to assist rather than replace a healthcare professional[20]. Some believed AI tools may be able to facilitate more person-centred care, by reducing the administrative burden on clinicians and allowing them to engage more with patients during appointments. Research has suggested that AI-generated patient summaries may be as acceptable as those written by junior doctors[36], though researchers have cautioned that AI summaries come with substantial risks, as omissions and inaccuracies may influence clinician decision making and lead to clinical errors[37,38].

Despite generally positive attitude towards AI-assisted SMRs, participants also expressed concerns regarding the potential impact of AI tools on patient safety and data security. These concerns highlight the importance of establishing appropriate regulatory frameworks for the use of AI in healthcare[39,40], as well as ensuring that AI tools are rigorously validated before being implemented in clinical practice[41]. Participants also expressed differing preferences regarding the disclosure of AI use, suggesting that expectations around how AI is communicated and explained may vary among PLwD and their carers. Participants further emphasised the importance of meaningfully involving PLwD and their carers in the design and development of AI tools to ensure they are relevant, acceptable, and responsive to the needs of this population [42].

### Strengths and Limitations

A strength of this study was the inclusion of participants with varied demographic and clinical characteristics, including individuals from different regions of the UK, a range of dementia diagnoses, and some minority ethnic backgrounds, which facilitated the exploration of diverse perspectives on the use of AI in SMRs[43].

Several limitations should be considered when interpreting these findings. First, although interviews focused on the use of AI in SMRs, some participants may have drawn on broader experiences of its use in medication reviews/healthcare when discussing the topic. Second, though the interviewer strongly emphasised that all opinions were welcome, it is possible that social desirability influenced the participant’s positive responses about using AI in healthcare. Third, the relatively younger age profile of carers and a male dominant sample may have resulted in higher AI literacy levels and more positive attitudes toward AI, as perceptions of digital technologies may vary by age and gender[44]. However, as data were analysed at the group level, the influence of individual characteristics on attitudes toward AI could not be assessed. Despite this, several participants expressed that they did not have a strong understanding of AI or technology and, therefore, could not go into detail about their views. Some of these participants may not have expressed concerns about AI because they were unaware of the known limitations of generative AI, including hallucinations and environmental impact[45,46]. Therefore, it is unclear to what extent positive attitudes towards AI were driven by a lack of awareness about AI, or whether they were willing to overlook these limitations due to the potential for positive impact on their healthcare.

### Conclusion

This study is the first to explore the attitudes of PLwD and their carers towards the use of AI in SMRs. Many participants viewed AI as acceptable, particularly where it might save clinician time and improve the quality of SMRs experiences. However, participants also highlighted the need to carefully consider patient safety, data security, the preservation of human interaction, and the implications of AI for clinicians’ decision-making and professional roles before implementing AI tools in clinical practice. These findings could be useful for informing the design of AI tools for dementia care by identifying the priorities and concerns of PLwD and carers.

## Acknowledgements

We thank the participants who generously invested their time and shared their experiences in interviews. We are also grateful to the healthcare professionals who attended workshops that informed the development of the interview guide for this study and the lived experiences of Patient and Public Involvement and Engagement (PPIE) representatives for their contributions to this project.

## Author Contributions

NT, SS, JG, and JD conceptualised and acquired the funding for the project. Study design was supervised by NT, SS, JG, and JD. BL carried out and transcribed interviews. Focus groups were carried out and transcribed by BL, and co-facilitated by either JD or JG. BL carried out data analysis, supervised by JD and JG. BL drafted the manuscript. JD led the later stages of manuscript revision, with contributions from NT, BL, JG, and SS. All authors critically reviewed the manuscript, provided feedback, and approved the final version.

## Data availability statement

Interview transcripts from the current study will not be made publicly available as this could compromise the participant’s privacy.

