## Supplementary File 1 for "Attitudes of People Living with Dementia and their Carers towards the use of Generative Artificial Intelligence to inform Structured Medication Reviews"

**AIMed Interview Guide for Focus Groups and Individual Interviews**

Please note that the questions provided are meant to serve as a guideline of the topics that will be discussed, and do not represent the actual wording of the questions asked in the interview. Questions and wording must be adjusted based on what is appropriate for that participant. The questions may also vary depending on the participants in each interview and the direction of the discussion. Additional probing questions will be asked as needed.

**Demographic data**

The following demographic data will be collected at the start of the interview/focus group:

- Age group (18-30, 31-40, 41-50, 51-60, 61-70, 71-80, 80+)
- Gender
- Role / Reason for participating (e.g. carer of someone with dementia/ person living with dementia)
- Ethnic background
- Geographical area of residence

Participants can decline to answer any of these demographic questions if they would prefer not to answer. Their response will be reported as ‘prefer not to say’.

The following ground rules for the focus group will be discussed at their start with the wider participants:

- Respect for All Participants
- Confidentiality
- Open Participation
- One Speaker at a Time
- Be Honest and Open
- Stay on Topic
- No Judgment
- Speak for Yourself
- Respect Time
- Limit Technology Distractions
- Participants will be encouraged to consider making changes to the ground rules and add any accommodations that would help them to feel comfortable.

**Topic guide for interviews and focus group**

1. *Concerns about medication management*

Explore the participants’ experience of medication reviews with healthcare professionals.

Prompts:

- Any concerns they have about taking many medications (e.g. challenges around taking and managing their medication, concerns about side effects or health, etc.)
- Who do you turn to if you have difficulties?
- Thoughts about their experiences talking/discussing with healthcare professionals (e.g. your hospital doctor, GP, pharmacist) about medication
- How often do you have SMRs? (Explain what a medication review is, if needed)
- Are you satisfied with your SMRs? Any thoughts about how medication reviews could be improved (Explain what a medication review is, if needed)

2. *Computer System and Attitudes to IT*

First, explore what do individuals/carers think about the potential uses of the computer system in terms of their medication.

Provide a brief explanation of what a computer could do to help professionals and, ultimately, people living with dementia and carers.

Prompts:

- What are the anticipated benefits of the computer system?
- [When developing the model] what should the computer system consider when *help*ing **professionals make decisions** about individuals’ medication lists (safety, healthcare records, people’s preferences)? (emphasize what should be included in the model rather than testing the model)

Explore participant attitudes towards IT and the use of the computer system.

- What are the main concerns and priorities in regard to using this sort of technology in healthcare? (e.g. mistakes, security concerns, etc.)
- How accepted would such a computer system be by individuals with dementia and their carers?
- Perceptions about using computer systems to help making decisions about medication for people living with dementia
  - Use of digital technology and artificial intelligence in healthcare more generally

A summary of the main points of discussion will be provided at the end of the interview/focus group and the next steps of the research project explained. We will thank individuals with dementia and carers for their time and contribution.
